# Rationale and study protocol of the MAMELI Cohort study (MApping the Methylation of repetitive elements to track the Exposome effects on health: the city of Legnano as a LIving lab)

**DOI:** 10.1101/2025.06.02.25328837

**Authors:** Valentina Bollati, Federica Rota, Laura Dioni, Chiara Favero, Simona Iodice, Marta Gallazzi, Andrea Spinazzè, Giacomo Fanti, Davide Campagnolo, Tiago Nardi, Davide Biganzoli, Eva Dariol, Rachele Matsagani, Mirjam Hoxha, Paola Monti, Benedetta Albetti, Letizia Tarantini, Davide Barbuto, Ester Luconi, Giuseppe Marano, Giacomo Biganzoli, Roberta Biciuffi, Patrizia Boracchi, Domenico M. Cavallo, Pilar M. Guerrieri, Fabio Mosca, Stefano Gustincich, Giovanni Sanesi, Luca Ferrari, Michele Carugno, Silvia Fustinoni, Angela Cecilia Pesatori, Luca Pandolfini, Michele Miragoli, Andrea Cattaneo, Elia Biganzoli

**Affiliations:** EPIGET Lab -Department of Clinical Sciences and Community Health, “Dipartimento di Eccellenza 2023-2027”, Università degli Studi di Milano, Milan, Italy; INES (Institute of Epigenetics for Smiles), Università degli Studi di Milano, Milan, Italy; Department of Science and High Technology, Università degli Studi dell’Insubria, Como, Italy; Unit of Medical Statistics, Department of Biomedical and Clinical Sciences (DIBIC), LITA Vialba campus, Università degli Studi di Milano, Milan, Italy; “Dipartimento di Architettura e Studi Urbani”, Politecnico di Milano, Milan, Italy; Università degli Studi di Milano, Milan, Italy; Non-coding RNAs and RNA-based Therapeutics, Istituto Italiano di Tecnologia (IIT), Genova, Italy; Department of Soil, Plant and Food Sciences, University of Bari Aldo Moro, Bari, Italy; Toxicology lab, Fondazione IRCCS Ca’ Granda Ospedale Maggiore Policlinico, Milan, Italy; Dipartimento di Medicina e Chirurgia, TecMed Lab, Università di Parma, Parma, Italy

**Keywords:** Repetitive elements, Exposome, DNA methylation, Epigenetics, Urban Health, Environmental wellbeing, Epidemiology

## Abstract

**Background:** The concept of the exposome encompasses all the factors influencing human health throughout the life course. The exposome induces epigenetic changes, such as DNA methylation, which influence gene expression and impact overall health. Several recent studies have explored how repetitive elements (REs) in the genome can be activated in response to environmental stimuli. However, most of these investigations have assumed that altered RE methylation is always detrimental to individual health. The MAMELI project proposes an alternative hypothesis: that some REs are plastic entities capable of responding physiologically to environmental stimuli without compromising genome stability. This hypothesis suggests that the ability of DNA to adapt to environmental triggers could be monitored and used as an indicator of health resilience.

**Methods:** To test this hypothesis, the MAMELI project will enroll 6,200 participants from the city of Legnano (Italy) and will be conducted in three main phases: i) A total of 200 healthy participants will undergo DNA methylation analysis through third-generation sequencing at two time points: T0 (baseline) and T1 (6 months after T0). This phase aims to identify a set of REs (“differential REs”) whose methylation changes in response to the exposome without affecting genome stability; ii) This phase will include 2,700 subjects (the original 200 participants from the discovery phase plus 2,500 additional subjects). The goal will be to develop a predictive algorithm (the MAMELI algorithm) that links the exposome to RE methylation status, creating a “RE methylation signature” reflecting the environmental impact on DNA methylation; iii) In this phase, the MAMELI algorithm will be applied to a separate cohort of 3,500 participants to compare the measured RE methylation signature with values predicted by the algorithm. Additionally, an intervention study will be embedded within the cohort to assess the reversibility of RE methylation following lifestyle changes.

**Discussion:** The MAMELI project offers a novel perspective in the field of epigenetics and environmental health, demonstrating how the epigenome can act as a sensor for environmental changes and how this interaction can be harnessed for disease prevention. If validated, the MAMELI algorithm could become a powerful tool for identifying individuals at risk and developing personalized interventions to improve global health outcomes.

## 1. Background

According to the WHO (World Health Organization), an estimated 24% of the global disease burden and 23% of all deaths can be attributed to modifiable environmental factors. Nine million deaths per year (16% of deaths worldwide) were attributed to air, water, and soil pollution alone [1].

Environmental threats to human health include numerous environmental pollutants including metals, phytoestrogens, polycyclic aromatic hydrocarbons, dioxin-like chemicals, polychlorinated biphenyls, phthalates, and several classes of pesticides.

Disease often results from interactions between genes and the environment experienced throughout the life course. Environmental exposures, however, cannot be limited to the above-mentioned toxicants, as it is becoming increasingly clear that non-conventional exposures (e.g., stressful events, social life, and parental care) can modulate disease risk.

The awareness that a multitude of factors may synergistically modulate the state of health/disease led to the concept of “exposome,” first defined in 2005 by Chris Wild [2]. The exposome includes the totality of human disease determinants encountered during the life course, and can be divided into three main domains: i) the internal environment (e.g., metabolism, endogenous circulating hormones, morphology, physical activity, gut microflora, inflammation, lipid peroxidation, oxidative stress, and aging); ii) specific external exposures (e.g., radiation, infectious agents, chemical contaminants and environmental pollutants, diet, lifestyle factors, tobacco, alcohol, occupation, and medical interventions); iii) the general external exposome that includes the wider social, economic, and psychological influences on the individual (e.g. social capital, education, financial status, psychological and mental stress, urban-rural environment, and climate) [3].

The aim of the exposomic concept would be to track an individual’s exposures (from pre-conception to death) to elucidate how exposures from our environment, diet, and lifestyle interact with our genetic backgrounds. There are no standard methods to fully characterize the exposome. The impacts of particular factors are affected by their close interconnections, while their comprehensive health effects are determined by their interactions. Analytical techniques are improving daily, but the analysis of so many factors in a large number of subjects is still too challenging. Moreover, the comprehensive follow-up of a human cohort over an entire lifespan is very difficult to execute.

Even if our capability to fully track and disentangle all exposomic information is limited, our DNA can record every stimulus that our bodies encounter throughout the life course. A growing compendium of evidence indicates that the exposome might induce epigenetic modifications, in particular, modifications of the DNA methylation pattern. Although the relative contribution of epigenetics to disease development is only partially understood, environmental epigenetics (i.e., the epigenetic pattern shaped by environmental exposures) holds great potential to provide a biomarker that may easily assess and summarize all lifestyle habits and environmental exposure effects.

Epigenetic mechanisms are flexible processes that change genome function under exogenous and endogenous influences, and also allow the stable propagation of gene activity states from one generation of cells to the next [4]. DNA methylation covalently modifies cytosine to form 5-methylcytosine (5mC), which represents 2–5% of all cytosines in mammalian genomes and is found primarily on CpG dinucleotides [5]. Studies of active DNA demethylation disproved the original assumption that DNA methylation is a stable epigenetic modification. Conversely, a vital aspect of DNA methylation is its dynamic and reversible nature, profoundly shaped by environmental stimuli.

If on the one hand, previous studies have had the great merit of demonstrating the link between exposures and modification of DNA methylation, on the other hand, they have been either too narrowly focused on specific pathogenetic mechanisms or, on the contrary, too broad (e.g., genome-wide associations) and consequently difficult to interpret.

Besides its well-known role in altering gene expression [6], DNA methylation also has an important function in the maintenance of genome integrity. This role is executed primarily through the methylation of repetitive DNA sequences and endogenous transposons that constitute approximately 40% of the human genome. These repetitive elements (REs) can be divided into two groups based on the presence or absence of long terminal repeats (LTRs). Non-LTR elements include short interspersed nuclear elements (SINEs, e.g., Alu) and long interspersed nuclear elements (LINEs) [7]. The majority of LTR elements are derived from human endogenous retroviruses and represent about 8% of the human genome [8,9]. The potential impact of altered RE methylation is underscored by the fact that REs, including LTR elements, constitute up to 40% of the human genome and can be activated under certain conditions [10,11]. Notably, increasing evidence shows that REs are frequently hypomethylated in various human pathologies, including cancer, cardiovascular and respiratory diseases, and psychiatric disorders. Historically, REs have been considered as “junk” DNA [12]. However, the high proportion of these sequences in our genome and their ability to respond to stimuli demonstrate the need for a paradigm shift to interpret current scientific evidence to understand the functions of REs and how they promote disease development [13].

## 2. Rationale

Recent studies have highlighted the complex interplay between environmental exposures and human health, yet the molecular mechanisms driving adaptability to external triggers remain poorly understood. While only 1-2% of the genome consists of protein-coding genes, the remaining non-coding regions, particularly repetitive transposable elements, comprise a significant portion of the genome and are now recognized as potential regulators of genomic stability. Traditionally labeled as “junk DNA,” REs are primarily epigenetically silenced by DNA methylation. Previous research has focused on the detrimental effects of altered RE methylation, often linking it to disease. However, this view overlooks the possibility that REs could function as dynamic elements that mediate physiological adaptability to environmental inputs, encapsulated in the concept of the exposome, the cumulative impact of all internal and external factors over an individual’s lifetime.

The current literature has not yet explored whether RE methylation could act as a beneficial mechanism for genomic adaptability to environmental stimuli. The MAMELI project seeks to address this gap by hypothesizing that REs are key elements in tracking DNA’s adaptability to such environmental triggers (Figure 1).

**Figure 1:**
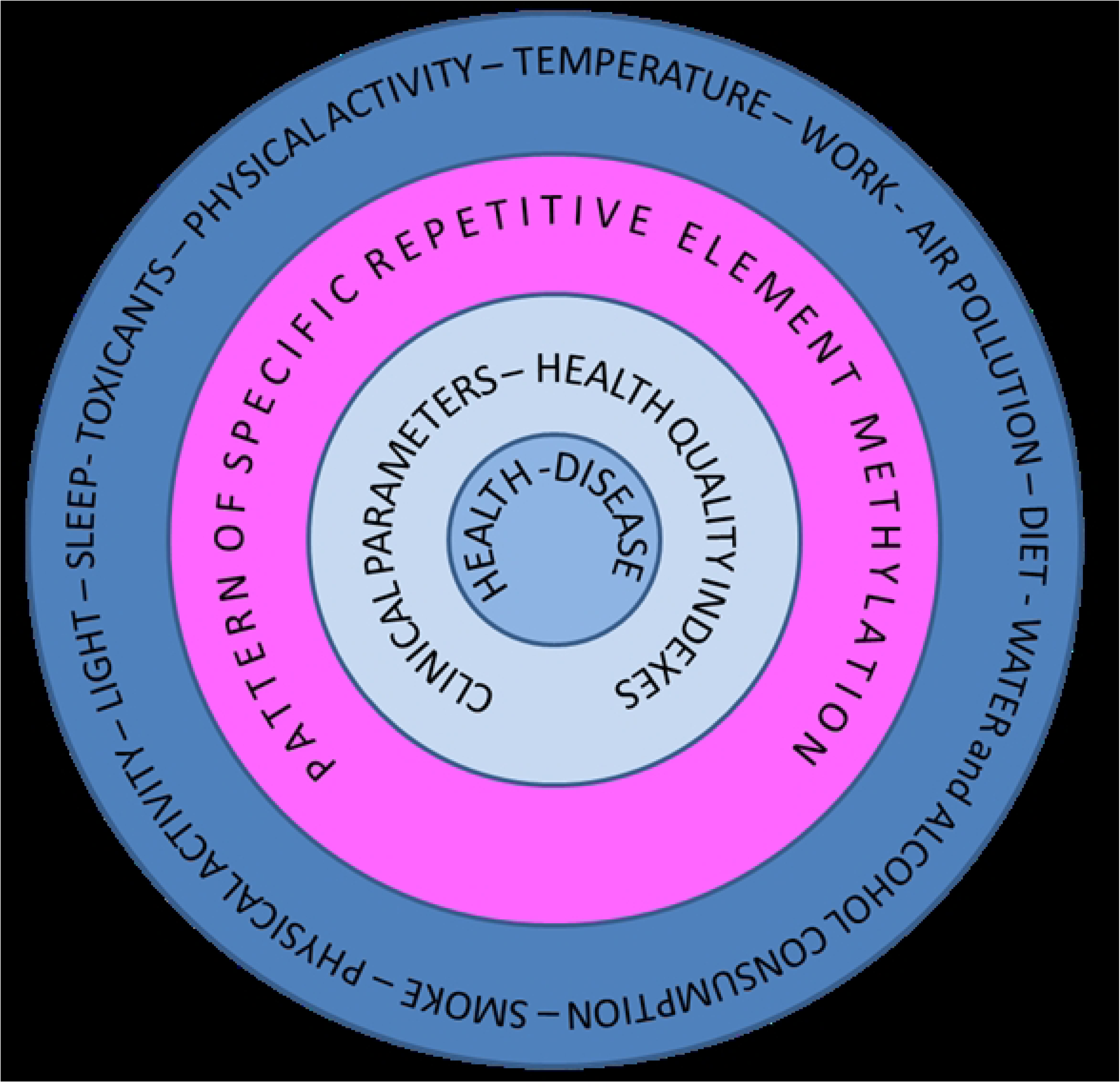
Conceptual model of the MAMELI Project

According to this hypothesis, REs act as highly sensitive epigenetic switches interspersed throughout the genome, typically repressed but capable of activation in response to external stimuli. This activation could induce genomic alterations or responses at multiple levels, facilitating an adaptive response to environmental challenges. However, global RE activation would likely lead to genomic instability, so we propose the existence of specific RE hotspots, methylated regions capable of sensing environmental cues and initiating an adaptive cascade. A healthy genome would react promptly to these inputs, while failure to do so could signal the genome’s inability to adapt, representing the first step toward pathogenesis.

By investigating this hypothesis, the MAMELI project will provide a novel perspective on the role of REs as mediators of genomic adaptability, offering a new understanding of how epigenetic mechanisms contribute to both health and disease in response to environmental pressures (Figure 2).

**Figure 2:**
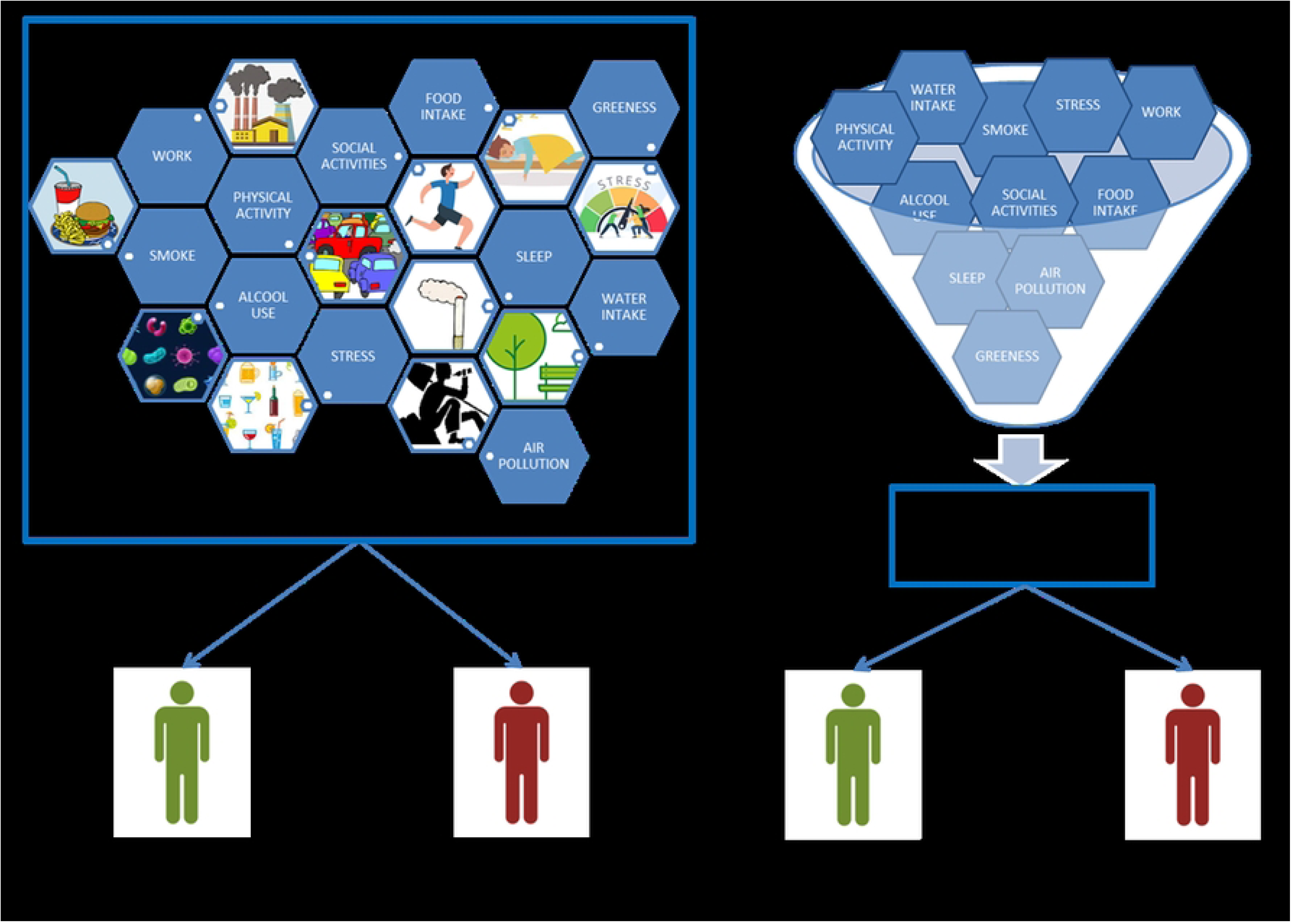
Exposome traditional approach versus what MAMELI is going to pursue.

## 3. Objectives and Research Questions

The MAMELI project aims to enroll 6,200 participants from Legnano (Milan), Italy, in a multi-tiered study, proceeding through three distinct investigative phases:

**Discovery Phase**: This initial phase involves 200 participants, each providing biological samples at two time points: baseline (T0) and six months later (T1). Using Nanopore direct genomic DNA sequencing, this phase will generate an unbiased screening of RE methylation status. By identifying REs with methylation patterns that can change without compromising genome stability, the discovery phase will set the groundwork for further analysis and classification of these methylation patterns.

**Tuning Phase**: In the second phase, a subset of 2,500 participants from the cohort will be analyzed to develop a predictive algorithm, referred to as the “MAMELI algorithm.” This algorithm will examine the relationship between RE methylation status and specific exposome profiles, building predictive RE methylation models based on the observed exposome patterns.

**Validation Phase**: In this final phase, the MAMELI algorithm will be applied to an additional 3,500 participants to compare the observed RE methylation signatures with the algorithm’s predictions. Additionally, an intervention study will be embedded within this phase to explore the reversibility of RE methylation status in response to lifestyle modifications, assessing their potential for health promoting/ disease preventive effects.

Throughout all three phases, data on exposome measures, clinical parameters, and health scores will be collected using wearable devices and a specially developed mobile app, as outlined in the Methods/Design section.

### Primary Objective

The primary objective of the MAMELI project is to investigate whether repetitive transposable elements (REs) act as dynamic epigenetic regulators that mediate human adaptability to the exposome. Specifically, the study aims to identify a methylation signature of REs that responds to environmental stimuli without compromising genome stability, and to develop a predictive algorithm (“MAMELI algorithm”) that models the relationship between RE methylation and specific exposome factors.

### Secondary Objectives

To validate the RE methylation signatures identified in the discovery phase across a larger cohort and determine their association with health vs disease profiles.

To assess the potential reversibility of RE methylation status following lifestyle modifications and examine the preventive potential of these changes on health outcomes.

To explore the long-term implications of RE methylation adaptability, focusing on chronic disease risk, hospitalizations, and mortality.

### Research Questions and Hypotheses

#### Discovery Phase

Research Question 1.1: Can we identify a pool of REs (“Differential REs”) whose methylation status changes in response to exposome stimuli without affecting genome stability?

Hypothesis 1.1: There exists specific REs whose methylation status is dynamically regulated by environmental factors and these changes can be detected using Nanopore sequencing in peripheral blood leukocytes over time.

#### Tuning Phase

Research Question 2.1: Which REs among the Differential REs are consistently modified by the majority of exposome factors?

Hypothesis 2.1: A subset of Differential REs will form a stable methylation signature that is influenced by a range of exposome factors and can be identified through targeted sequencing.

Research Question 2.2: Can we develop a predictive algorithm (MAMELI algorithm) that models the relationship between specific exposome patterns and the RE methylation signature?

Hypothesis 2.2: The MAMELI algorithm will accurately predict changes in the RE methylation signature based on the exposome data collected.

#### Validation Phase

Research Question 3.1: Does the RE methylation signature identified in the discovery phase accurately reflect the exposome in the broader cohort?

Hypothesis 3.1: The RE methylation signature will correlate with exposome factors in the larger study cohort, supporting its use as a biomarker of environmental adaptability.

Research Question 3.2: How does the difference between the measured RE methylation signature and the values predicted by the MAMELI algorithm reflect the individual’s adaptability to environmental stimuli?

Hypothesis 3.2: A greater difference between the observed and predicted methylation signatures will indicate a reduced ability to adapt to external stimuli, potentially serving as an early marker of disease risk.

Research Question 3.3: Is there an association between the difference in RE methylation signature (as per Aim 3.2) and clinical parameters or health scores recorded by wearables and the mobile app?

Hypothesis 3.3: A significant association will be found between the deviation in methylation signature and poor health outcomes, as reflected in clinical parameters and wearable data.

Research Question 3.4: Can lifestyle modifications reverse the RE methylation changes and improve health outcomes?

Hypothesis 3.4: Intensive lifestyle interventions will positively influence RE methylation signatures and health outcomes, supporting their role in preventing disease progression.

### Long-Term Objective

Research Question 4.1: What is the long-term association between RE methylation adaptability and future health outcomes, including chronic diseases, hospitalizations, and mortality?

Hypothesis 4.1: Subjects with greater discrepancies between predicted and observed RE methylation signatures will exhibit higher risks for chronic diseases and adverse health outcomes over time.

## 4. Methods/Design Study Design

The MAMELI project is a longitudinal cohort study designed to explore the role of repetitive transposable elements (REs) in mediating human adaptability to environmental factors, collectively known as the exposome. The study will involve three phases: discovery, tuning, and validation, with a final interventional component embedded in the cohort. The primary objective is to identify RE methylation signatures that respond to environmental stimuli and to develop the MAMELI algorithm for predicting health outcomes based on these signatures (Figure 3).

**Figure 3:**
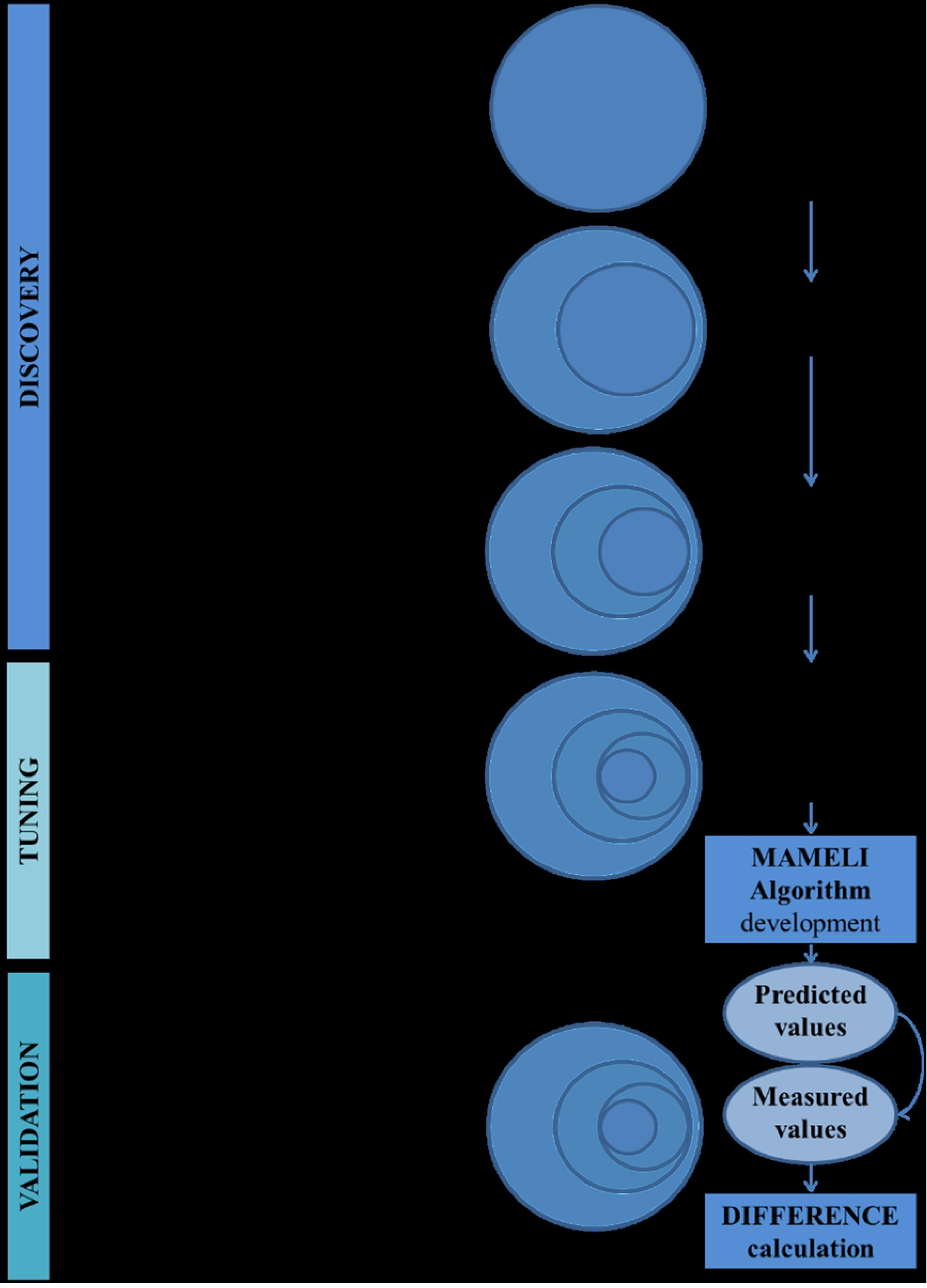
Process followed to develop and validate the MAMELI algorithm.

### Setting

The study will be conducted in Legnano, a city in the Lombardy Region of Italy, with approximately 60,000 residents. Legnano offers a strategic location due to its proximity to Milan, manageable population size, and location within the Po Valley, a highly polluted area (Figure 4). The study is supported by the local government and will utilize local infrastructure for recruitment and sample collection.

**Figure 4:**
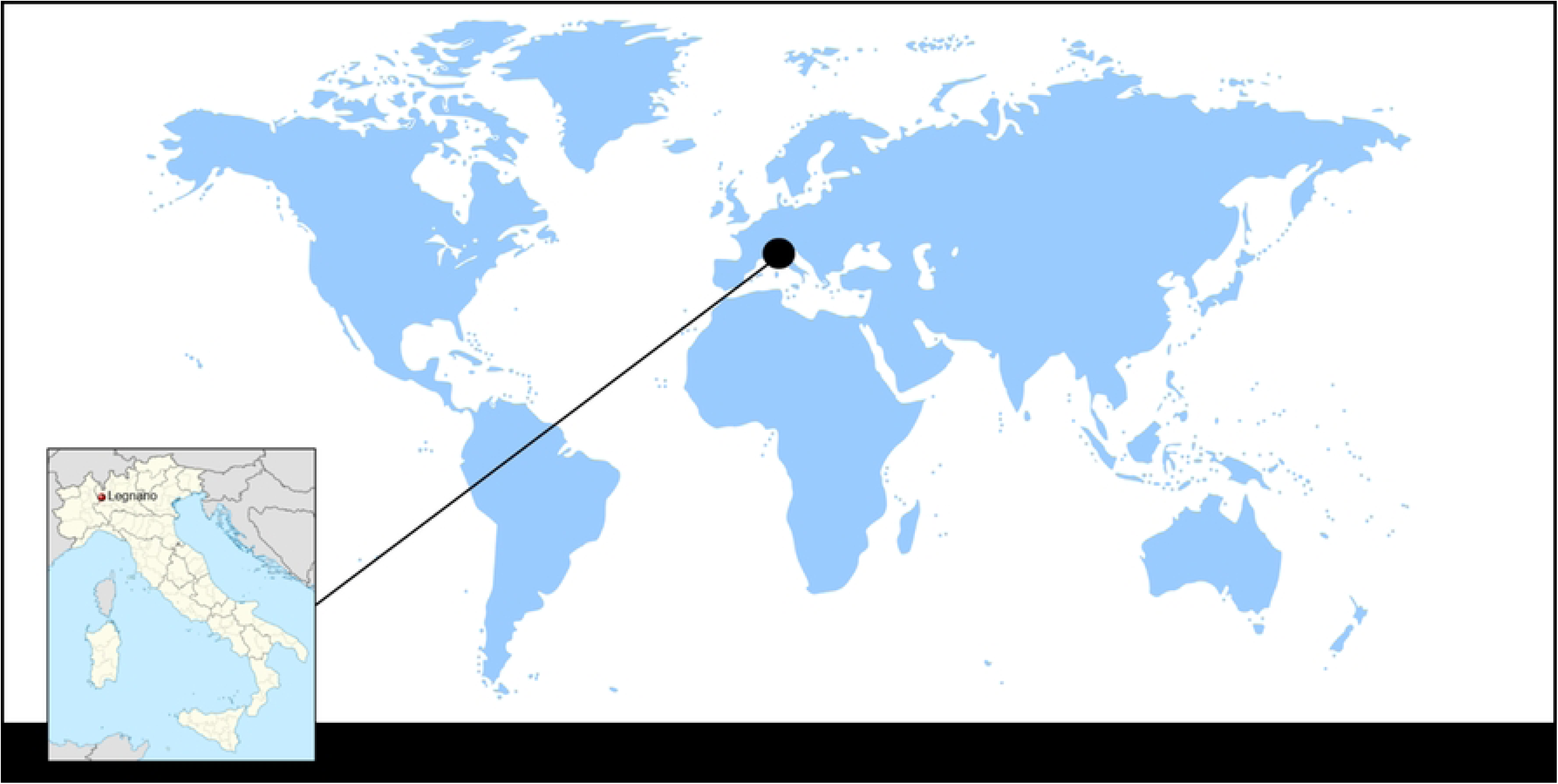
Map showing the location of Legnano, home to the MAMELI project. Positioned in the Lombardy region of northern Italy, Legnano is strategically located near Milan, enhancing its accessibility and relevance within the project’s framework.

### Participants

The study will enroll 6200 adult participants (aged >18 years) from the city of Legnano, divided into three phases:

Discovery Set: 200 participants who will provide biological samples at two time points (T0 at baseline and T1 at 6 months).

Tuning Set: 2500 participants who will provide samples at T0 only. Validation Set: 3500 participants who will provide samples at T0 only. **Recruitment start date:** 3 January 2024

**Expected recruitment end date:** 2 January 2027

### Inclusion Criteria

Adult participants (age >18) Residents of Legnano **Exclusion Criteria:**

Impairments precluding informed consent Cancer within the last 2 years (Discovery set only)

Uncontrolled dysrhythmias, heart failure, valvular or myopericardial disease (Discovery set only) Major chronic diseases (Discovery set only)

Participants will be recruited through community outreach, including a city-wide awareness day and ongoing recruitment at a central city location, supported by local blood donation centers (AVIS Legnano). Signed informed consent will be obtained from all participants.

### Intervention

An intervention will be conducted for participants who show a significant mismatch (>5%) between the RE methylation signature predicted by the MAMELI algorithm and the observed values. These individuals will be offered an intensive lifestyle modification program, including dietary changes, increased physical activity, and psychological well-being support. Participants who refuse the intervention will receive standard clinical care. Outcomes will be compared between the intervention and control groups based on RE methylation and health outcomes.

### Outcomes

#### Primary Outcomes

Identification of RE methylation signatures that change in response to exposome stimuli without destabilizing the genome.

Development and validation of the MAMELI algorithm to predict RE methylation changes based on exposome data.

#### Secondary Outcomes

Association between RE methylation signatures and health parameters (e.g., heart rate variability, sleep quality, physical activity).

Reversibility of RE methylation changes following lifestyle intervention and their impact on long-term health outcomes.

### Measurements

#### Exposome and Clinical Parameter Measurements

To comprehensively characterize the participants’ exposomes and collect clinical parameters, the MAMELI project utilizes a combination of wearable devices, a custom-developed mobile app, geographical information system (GIS) data, and high-throughput targeted biomonitoring of biological samples.

#### Wearables

Participants wear a commercially available high-end wristband capable of monitoring a range of health and fitness parameters. The device continuously records oxygen saturation, skin temperature, breathing rate, heart rate variability (HRV), resting heart rate, sleep quality, and physical activity by integrated accellerometers and photoplethysmography. Additionally, the wristband monitors environmental exposures. Data from the wearable devices will be integrated into artificial intelligence (AI) models to support the analysis of health trends and responses to environmental exposures.

#### Mobile App Development

The MAMELI-App, specifically developed for this study, tracks participants’ dietary habits and mood. For two weeks preceding sample collection (T0 and possibly T1), participants carry smartphones with the MAMELI-App installed. To capture dietary intake, participants will photograph their meals (which will be then used for image recognition to identify food items). Mood will be assessed through simple, self-reported queries about the participants’ feelings.

#### Air Pollution Monitoring and Exposure Assessment

To assess participants’ exposure to key airborne pollutants, PM2.5 (particulate matter <2.5 micrometers), ozone (O₃), and nitrogen dioxide (NO₂), an integrated approach combining land use regression (LUR), chemical transport models (CTM), and geographic information system (GIS) data will be utilized.

A Wireless Sensor Network (WSN) consisting of 16 low-cost air quality monitors has been established across Legnano. The placement of these monitors was strategically designed to cover a range of zones, including urban, suburban, traffic, and industrial areas, while ensuring broad spatial coverage. Additionally, technical factors such as ground-level positioning, power supply, and Wi-Fi connectivity were considered during deployment. Since the city of Milan is the main work destination for commuters living in Legnano, an additional low-cost monitor was installed in Milan downtown to collect real-time data also in this location. The devices employed are BlueSky™ 8145 sensor system (TSI Inc., Minnesota, USA), which offer real-time measurements with a 1-minute resolution. These monitors measure the target pollutants (PM2.5, NO₂, O₃) along with other pollutants (CO₂, CO, SO₂) and meteorological parameters (temperature, relative humidity (RH)) with high reliability [14–17]. To ensure data accuracy and facilitate quality control, three reference-grade instruments compliant with EN/ISO standards have been installed at a selected urban background location, alongside a co-located BlueSky™ monitor. These instruments include:

- DPA14 Gravimetric Sampler (XEarPro Srl, Monza e Brianza, Italy) – Provides accurate PM2.5 concentrations with a 24-hour time-integrated average.
- AC32e NO₂ Monitor (ENVEA Inc., Poissy Cedex, France) – Measures NO₂ concentrations at a 1-minute resolution.
- O342e O₃ Monitor (ENVEA Inc., Poissy Cedex, France) – Provides O₃ concentrations with 1-minute resolution.

The data from these reference instruments will serve as a calibration standard, ensuring the reliability of measurements obtained from the low-cost sensors in the WSN. These reference measurements will also enable the correction of sensor-derived data, improving the accuracy of the study’s exposure assessments. As participants are likely to travel beyond Legnano’s boundaries, additional data will be incorporated from the Lombardy Regional Environmental Protection Agency (ARPA Lombardia). Hourly pollutant data from regional monitoring stations will complement the WSN measurements. Missing values in this dataset will be addressed through an imputation algorithm that combines annual averages and readings from the nearest correlated monitoring stations.

To achieve precise, location-based exposure estimates, participants’ GPS data will be utilized. This approach ensures accurate tracking of individual movements and pollutant exposure, both within and outside the study area.

### Greenness

We adopted different procedures for assessing the greenness exposure. We calculated green and grey spaces surrounding the residential address of each participant based on the average of 2D and 3D indicators of green and grey spaces across buffers of 50 m, 100 m, 300 m, and 500 m around everyone’s geocoded residential address [18]. For the 2D approach we calculated of the NDVI using high-resolution RapidEye data that encompasses a 5-satellites constellation each one carrying a multispectral sensor acquiring data in five spectral bands (blue: 440–510 nm; green: 520–590 nm; red: 630–685 nm; red edge: 690–730 nm; and near infrared: 760–850 nm) at approximately 5 m x 5 m resolution (Planet Labs, 2016). For the 3D approach we used light detection and ranging (LiDAR) data to develop our 3D indicators of green and grey spaces, including green volume, grey volume, Normalized Difference of Green-Grey Volume (NDGG) descripted by Giannico et al. [19] and Spano et al. [20] in previous research. We also derived tree count from LiDAR data analysis. LiDAR data was provided by the “Città Metropolitana di Milano”. Data were acquired in 2022 with a resolution of about 16 points per m^2^.

### Urban Environmental Analysis

The study involves urban analysis, both historical and present, in order to provide an overall deeper understanding of the impact of the environment on individuals who are daily exposed. Indeed, since the choices of the past have shaped Legnano today, reconstructing key historical events, such as its industrial legacy, has been crucial to gain critical insights on critical issues, strength, and planning future actions. Whereas the urban analysis in the present, consisting in urban mappings of people’s life habits collected through detailed individual GPS tracking, provides charts and statistics on how people actually use and interact with the city, i.e. if they are spending most of their time in town or mostly living as commuters, if they commute with cars, public transports or using bicycles, how much time they are in beneficial green areas or stuck in traffic, etc. These urban lifestyle habits are tightly connected and inevitably affect, positively or negatively, people’s quality of life and wellbeing. “The Urban Exposome,” which has proven to be one of the most important factors capable of influencing people’s lives, has been addressed in the study in order to provide evidence and explanation on how the way people use the city changes not just the quality of life but also life expectancy, producing direct effects on DNA methylation.

### High-Throughput Targeted Biomonitoring

Liquid chromatography-tandem mass spectrometry (LC-MS/MS) will be employed for the targeted biomonitoring of chemical pollutants in urine and plasma samples. The pollutants measured will include:

- Phthalates, including di-2-ethylhexyl terephthalate (DEHTP).
- Polycyclic aromatic hydrocarbons (PAHs): 9 metabolites.
- Per- and polyfluoroalkyl substances (PFASs): 30 legacy and emerging PFASs in plasma, such as HFPO-DA, DONA, and cC6O4.
- Tobacco smoke exposure markers: Urinary nicotine and cotinine as biomarkers of active and passive smoking.
- Urinary mercapturic acids: 17 metabolites of electrophilic intermediates from occupational and environmental toxicants.

Urinary metal concentrations will be quantified using inductively coupled plasma mass spectrometry (ICP-MS). The metals analyzed will include a wide range of trace elements such as aluminum, cadmium, lead, mercury, and nickel, among others.

### Third-Generation Sequencing for Methylation Analysis

Repetitive element (RE) 5-methylcytosine (5mC) methylation patterns are challenging to accurately characterize using traditional next-generation sequencing (NGS) technologies due to limitations in bioinformatic tools and the inherent complexity of repetitive sequences [21]. Whole-genome bisulfite sequencing (WGBS), which involves sodium bisulfite treatment to differentiate methylated from unmethylated cytosines, provides comprehensive methylation profiles with relatively high quantitative accuracy, despite being dependent on a complete and homogeneous chemical conversion efficiency across samples. However, the short-read length of NGS restrict the ability to precisely locate specific REs within the genome due to their repetitive nature, leading to ambiguous sequence alignment. For studies involving REs, the context of the methylation event is crucial, as the exact genomic positioning of each RE is necessary to identify specific methylation hotspots.

Third-generation sequencing offers a significant advantage for studying 5mC methylation and other epigenetic modifications without the need for bisulfite conversion. Nanopore sequencing can directly detect methylation on long DNA strands, providing greater genomic context and allowing for the precise mapping of RE methylation events. Furthermore, it is not dependent on an optimal efficiency of chemical conversion and eliminates the need for PCR amplification, which can introduce biases in methylation studies.

Given these advantages, third-generation sequencing will be employed in the discovery phase of the MAMELI study. This phase will involve an unbiased screening of RE methylation in 200 subjects at two time points (T0 and T1). The comprehensive nature of third-generation sequencing will allow for the identification of specific REs that exhibit modifiable methylation patterns in response to environmental stimuli. These findings will form the basis for more targeted methylation analyses in subsequent study phases.

The resulting data will be subjected to relatively straightforward statistical comparisons between T0 and T1 to identify REs that are modifiable, without compromising genomic stability. These modifiable REs will then be the focus of targeted methylation analysis in later stages of the study.

### Targeted Methylation Analysis

Targeted methylation analysis will be applied during the “tuning” and “validation” phases of the MAMELI project to focus on specific repetitive elements (REs) identified in the discovery phase. High-quality DNA samples will undergo bisulfite treatment, a process that converts unmethylated cytosines to uracil while leaving methylated cytosines unchanged. This enables the detection of methylation at cytosine-phosphate-guanine (CpG) sites by comparing the ratio of cytosines and thymines after amplification and sequencing.

A custom-designed targeted panel will be created to detect methylation at single genomic positions of the previously identified REs with high specificity. This approach will enrich for those REs found to be modifiable during the discovery phase, ensuring precise quantification of their methylation status. Methylation sequencing will be performed using the Next-Seq2000 Illumina platform, which generates detailed methylation data for each targeted RE.

The methylation data obtained from this targeted sequencing will be critical for the development and validation of the MAMELI algorithm, which aims to model the relationship between RE methylation and exposome factors.

### Statistical Analysis, Machine Learning, and MAMELI Algorithm Development

The MAMELI project will use a combination of descriptive statistics, multivariate analysis, and computational statistics/machine learning methods to develop and validate the MAMELI algorithm, which models the relationship between repetitive element (RE) methylation signatures and exposome factors.

### Descriptive Statistics

Descriptive statistics will be used to explore and summarize data from all study phases. The distribution of the main variables of interest will be examined graphically and resorting to density estimation methods and multivariate dependence structure detection. In addition advanced methods for mixed continuous and categorical data will be adopted like Multiple Correspondence Analysis or Generalized Principal Component Analysis (GPCA) while Generalized Canonical Correlation Analysis (GCCA) and its extension for dealing with data sparsity will be performed to explore the relationship between RE methylation and exposome variables. This initial analysis will provide an overview of the sample features and identify potential outliers or data anomalies.

### Statistical Analysis for the Discovery Phase

To identify differential RE methylation between T0 (baseline) and T1 (6 months), paired t-tests will be conducted on standardized measurements of methylation levels (|T0-T1|). Given the large number of comparisons across multiple RE sites, false discovery rate (FDR) control will be applied to adjust for multiple testing and reduce the likelihood of Type I errors. Beside the standard univariable paired t-tests, Multivariate Analysis of Variance for Repeated Measures (MANOVA-RM) and Functional Principal Component Analysis (fPCA) approaches will be adopted accounting for the correlation patterns of the methylation levels in the different RE sites.

Differential RE methylation will be ranked according to the magnitude of absolute differences, and results will be visualized using volcano plots. Boosting algorithms will be also applied to enhance detection of the most significant methylation changes.

### Multivariate Analysis for the Tuning Phase

In the tuning phase, multivariate regression analysis will be used to identify REs whose methylation is significantly impacted by a majority of exposome factors. These REs will form the “RE methylation signature,” a robust indicator of environmental response. Multivariate linear regression models will be used to model the dependence between the RE methylation signature and various exposome factors, while controlling for demographic and clinical characteristics. To this aim, Multivariate Adaptive Regression Splines (MARS), Functional Regression Models will be adopted in preliminary phase to explore the dependence relationships.

### Development of the MAMELI Algorithm

The RE methylation signature will be further investigated to develop the MAMELI predictive algorithm, which will estimate the relationship between exposome factors and RE methylation changes. Partial least squares (PLS) regression analysis will be employed to model this relationship. The predictive accuracy of the MAMELI algorithm will be assessed using bootstrap resampling techniques to ensure that the model is generalizable and robust across different subsets of the data.

### Validation Phase

In the validation phase, the MAMELI algorithm will be tested to evaluate its predictive accuracy. For each study subject, the difference between the measured RE methylation signature and the predicted values from the MAMELI algorithm will be calculated. This difference will serve as an indicator of an individual’s capacity to adapt to environmental stimuli [22].

The association between this difference and clinical parameters, as well as health scores (collected via wearables and the MAMELI app), will be assessed using multivariable generalized linear regression models. Additionally, in the interventional phase, the impact of lifestyle modifications on the RE methylation difference will be evaluated using generalized linear regression models for repeated measures.

### Long-Term Follow-Up

The study cohort will be followed longitudinally for several years, allowing multivariate analyses to investigate whether the RE methylation signature difference is associated with hard health outcomes, including the incidence of chronic diseases, hospitalizations, and mortality.

### Software and Implementation

All statistical analyses will be performed using R statistical packages (R Core Team, 2021), with specialized packages used for machine learning, multivariate analysis, and data visualization. The results will provide key insights into the relationship between RE methylation, exposome factors, and health outcomes, enabling the development of a predictive model for personalized health monitoring.

### Sample Size Calculation

The study aims to enroll 6200 participants, representing ∼10% of Legnano’s population. The sample size is based on statistical power calculations to detect significant differences in RE methylation between groups with an expected power of 80% and alpha of 0.05, accounting for multiple testing using false discovery rate (FDR) adjustments. The discovery phase involves 200 subjects, providing sufficient power to identify RE methylation changes over time. The tuning and validation sets (n=2500 and n=3500, respectively) allow for robust multivariate analysis of RE-exposome interactions and validation of the MAMELI algorithm.

## 5. Ethics and Dissemination

### Ethical Approval

This study has received ethical approval from the Ethics Committee of the University of Milan. The approval was granted on March 21st, 2023, with the reference number 35.23. All study procedures will be conducted in accordance with the ethical standards set forth by the committee, ensuring compliance with relevant guidelines and regulations for research involving human participants.

### Consent

Informed consent will be obtained from all participants prior to their inclusion in the study. Participants will be provided with detailed information about the study’s objectives, procedures, potential risks, and benefits. This information will be presented in clear, non-technical language both in written form and during a face-to-face discussion with a study team member. Participants will have the opportunity to ask questions and clarify any concerns before signing the consent form. The consent process will be conducted in compliance with ethical standards, ensuring that participation is entirely voluntary. Participants will be informed of their right to withdraw from the study at any time without any consequence. Digital and physical copies of the signed informed consent forms will be securely stored in compliance with the General Data Protection Regulation (GDPR) and local regulations on privacy and data protection.

### Dissemination Plan

The results of the MAMELI project will be disseminated through multiple channels to ensure broad reach among stakeholders, the academic community, and participants.

### Academic Community

Findings will be shared through peer-reviewed scientific journals, focusing on high-impact publications in the fields of epigenetics, environmental health, and epidemiology. The results will be presented at international and national scientific conferences, workshops, and symposia to foster knowledge exchange with other researchers and experts in related fields. Data and methodologies will be made available, where appropriate, for replication and further research by other scientists, promoting transparency and advancing the field.

### Stakeholders and Policy Makers

Reports and policy briefs will be created for public health officials, regulatory agencies, and other stakeholders to inform policy and decision-making related to environmental health and public health strategies. Regular updates and a final report will be shared with the local government of Legnano and other collaborating institutions to highlight the community impact of the study and its broader implications for public health interventions.

### Participants and the General Public

A summary of the findings, written in accessible, non-technical language, will be shared with participants through a dedicated section of the MAMELI project website (mameli.unimi.it), ensuring transparency and acknowledging their contribution to the study. In addition, to broaden engagement with the general public, MAMELI will host public outreach events and share regular updates on social media, encouraging discussions on environmental health and how epigenetics influences human adaptability. To reach a diverse and younger audience, MAMELI launched a dedicated Instagram page (https://www.instagram.com/mameli_2023/). Through this platform, followers can stay updated on project developments, activities, and initiatives, making it easy for the community to connect with the project and learn about cutting-edge research in an accessible, engaging way.

## 6. Discussion

### Strengths and Limitations

The MAMELI project boasts several key strengths. First, the study’s large sample size (6200 participants) enhances its statistical power and allows for robust multivariate analyses. The longitudinal cohort design, which includes three phases (discovery, tuning, and validation), ensures comprehensive exploration and validation of RE methylation signatures across a diverse population. Another strength is the integration of cutting-edge technologies such as third-generation nanopore sequencing and high-throughput targeted biomonitoring, providing precise and high-resolution methylation data. Additionally, the use of wearable devices and a custom-developed mobile app allows for real-time tracking of individual exposomes, giving a detailed view of environmental and lifestyle factors influencing health.

However, the study also has limitations. One potential limitation is the reliance on bisulfite treatment and targeted methylation sequencing, which, while offering specificity, might miss methylation changes in regions outside the targeted REs. Another challenge is the use of short-read sequencing (Illumina), which may limit the resolution of some repetitive elements due to their high-density regions. To address this, a targeted nanopore approach is considered for certain cases. Moreover, the study focuses primarily on a single geographic location (Legnano, Italy), which may limit the generalizability of the results to populations living in different environmental contexts. Additionally, while the wearable devices and mobile app provide real-time exposure data, there is a possibility of self-reporting bias, particularly in the app-based dietary and mood tracking components. More comprehensive urban analysis could be integrated considering that noise, temperature and other urban stressors may subtly influence the quality of life, wellbeing, livability and overall perceptions/metylation of the individuals. Lastly, despite the large sample size, lifestyle interventions may not capture the full complexity of long-term exposomic influences or account for individual variability in response to environmental stimuli.

### Relevance

The findings of the MAMELI project have significant potential to advance public health knowledge and practice. By identifying RE methylation signatures that respond to environmental exposures, this study could provide novel biomarkers for tracking how environmental and lifestyle factors influence health at the genomic level. These biomarkers could be used to develop predictive models for personalized health risk assessments, enabling early detection of maladaptive genomic responses and identifying individuals at risk of developing chronic diseases. Moreover, the development of the MAMELI algorithm offers a practical tool for modeling the complex interactions between the exposome and epigenomic changes. This predictive model could inform public health interventions by identifying critical environmental exposures that have the greatest impact on health, thereby helping policymakers prioritize regulations and preventive strategies. Additionally, the project’s focus on lifestyle interventions underscores the potential for mitigating adverse health outcomes through personalized preventive measures, contributing to the growing field of precision medicine. In the long term, the study’s findings could be applied to monitor population health trends in response to changing environmental conditions, such as pollution or climate change, further informing public health strategies at both the individual and population levels. Ultimately, the MAMELI project has the potential to shift the paradigm in environmental health by demonstrating the dynamic and adaptive nature of RE methylation in response to the exposome, providing new avenues for research and public health policy aimed at preventing disease and promoting healthy longevity.

## 7. Declarations

### Ethics approval and consent to participate

This study received ethical approval from the Ethics Committee of the University of Milan. The approval was granted on March 21st, 2023, with the reference number 35.23. All study procedures will be conducted in accordance with the ethical standards set forth by the committee, in line with relevant national and international guidelines for research involving human participants.

### Consent for publication

Not applicable

### Availability of Data and Materials

All data and materials generated in the course of the MAMELI project will be handled in accordance with ERC guidelines and the FAIR (Findable, Accessible, Interoperable, and Reusable) principles. Datasets, protocols, and relevant supplementary materials will be made publicly available via institutional repositories (e.g., Zenodo, OpenAIRE) upon project completion or as early as the project’s stages allow, depending on publication timelines and data sensitivity. Access to these materials will be granted under open-access licenses, ensuring broad dissemination while preserving the integrity of the research. Where applicable, datasets will be fully anonymized prior to deposition, in compliance with the General Data Protection Regulation (GDPR) and ethical best practices for research involving human subjects. Sensitive or potentially identifiable data will be carefully evaluated and handled under strict ethical protocols. All data management and dissemination procedures will adhere to the Do No Significant Harm (DNSH) principle, ensuring that the project does not negatively impact the environment or society during the collection, processing, or sharing of research materials. Specific details about the datasets and access conditions will be provided in each corresponding publication, enabling further research and replication efforts by the scientific community.

### Authors’ Contributions

VB, EB, FR conceived the study and developed the research design, BA, DB, ED, LD, MH, RM, PM, LT, LF, perform sample collection, DNA preparation and targeted methylation analyses; MG, ED, RM, collect on-site data; DB, TN, LP perform bioinformatic analyses; GB, PB, CF, SI, GM, EL, EB, perform statistical analyses, AC, DC, GF, AS, EL, EB, develop air pollution exposure modeling, PMG, FM, GS investigate the role of architecture, greenness and urban health; MC, Occupational Health Assessment; SF, Targeted Biomonitoring; LP, TN, LD, ED, SG perform third-generation sequencing analyses; MM, cardiac signal analyses; FR managed project logistics, including resource allocation and timeline coordination, VB secured funding for the project through the ERC Consolidator Grant, VB, EB, ACP supervised the research activities and provided oversight on all aspects of the project, VB, EB, FR drafted the manuscript and organized the main findings, BA, DB, DB, GB, PB, DC, ED, LD, GF, CF, MG, MH, SI, EL, GM, RM, PM, TN, AS, LT, PMG, FB, SG, GS, LF, MC, SF, ACP, LP, MM, AC reviewed and edited the manuscript, contributing to revisions and final approval.

## Data Availability

No datasets were generated or analysed during the current study. All relevant data from this study will be made available upon study completion.

## Acknowledgements

We would like to express our sincere gratitude to the following individuals and institutions for their invaluable contributions to this study: Thanks to the Administration of the Municipality of Legnano, particularly Anna Pavan and Simone Bosetti, for their support in the conceptualization and facilitation of the study; AVIS Legnano and AVIS Milano for their essential collaboration in carrying out the blood sample collections for the study; The AVIS Legnano volunteers, whose dedication and commitment were fundamental to the successful completion of this project; Samanta Rigo and Luca Beccato for their dedication in supporting subjects recruitment; Yeraldin Castillo De Spelorzi and Diego Vozzi of the Genomics Facility of the Istituto Italiano di Tecnologia (IIT) of Genova for support in long read sequencing activity; Laura Cantone, Anna Cerini, Manuela Marconi, Alessandro Gabrieli and Davide Bellavite for their support with the intense administrative procedures. Alice Posfortunato and Lidia Mantia for their assistance and guidance in navigating ethical procedures; All the volunteers of the MAMELI study, whose participation made this research possible.

